# Fine-tuned large language models for answering questions about full-text biomedical research studies

**DOI:** 10.1101/2024.10.28.24316263

**Authors:** Kaiming Tao, Jinru Zhou, Zachary A. Osman, Vineet Ahluwalia, Chiara Sabatti, Robert W. Shafer

## Abstract

**Background:** Few studies have explored the degree to which fine-tuning a large-language model (LLM) can improve its ability to answer a specific set of questions about a research study.

**Methods:** We created an instruction set comprising 250 marked-down studies of HIV drug resistance, 16 questions per study, answers to each question, and explanations for each answer. The questions were broadly relevant to studies of pathogenic human viruses including whether a study reported viral genetic sequences and the demographics and antiviral treatments of the persons from whom sequences were obtained. We fine-tuned GPT-4o-mini (GPT-4o), Llama3.1-8B-Instruct (Llama3.1-8B), and Llama3.1-70B-Instruct (Llama3.1-70B) using a quantized low rank adapter (QLoRA). We assessed the accuracy, precision, and recall of each base and fine-tuned model in answering the same questions on a test set comprising 120 different studies. Paired t-tests and Wilcoxon signed-rank tests were used to compare base models to one another, fine-tuned models to their respective base model, and the fine-tuned models to one another.

**Results:** Prior to fine-tuning, GPT-4o displayed significantly greater performance than both Llama3.1-70B and Llama3.1-8B due to its greater precision compared with Llama3.1-70B and greater precision and recall compared with Llama3.1-8B; there was no difference in performance between Llama3.1-70B and Llama3.1-8B. After fine-tuning, both GPT-4o and Llama3.1-70B, but not Llama3.1-8B, displayed significantly improved performance compared with their base models. The improved performance of GPT-4o resulted from a mean 6% increased precision and 9% increased recall; the improved performance of Llama3.1-70B resulted from a 15% increased precision. After fine-tuning, Llama3.1-70B significantly outperformed Llama3.1-8B but did not perform as well as the fine-tuned GPT-4o model which displayed superior recall.

**Conclusion:** Fine-tuning GPT-4o and Llama3.1-70B, but not the smaller Llama3.1-8B, led to marked improvement in answering specific questions about research papers. The process we describe will be useful to researchers studying other medical domains.

**AUTHOR SUMMARY:** Addressing key biomedical questions often requires systematically reviewing data from numerous studies—a process that demands time and expertise. Large language models (LLMs) have shown potential in screening papers and summarizing their content. However, few research groups have fine-tuned these models to enhance their performance in specialized biomedical domains. In this study, we fine-tuned three LLMs to answer questions about studies on the subject of HIV drug resistance including one proprietary LLM (GPT-4o-mini) and two open-source LLMs (Llama3.1-Instruct-70B and Llama 3.1-Instruct-8B). To fine-tune the models, we used an instruction set comprising 250 studies of HIV drug resistance and selected 16 questions covering whether studies included viral genetic sequences, patient demographics, and antiviral treatments. We then tested the models on 120 independent research studies. Our results showed that fine-tuning GPT-4o-mini and Llama3.1-Instruct-70B significantly improved their ability to answer domain-specific questions, while the smaller Llama3.1-Instruct-8B model was not improved. The process we described offers a roadmap for researchers in other fields and represents a step in our attempt towards developing an LLM capable of answering questions about research studies across a range of pathogenic human viruses.

## INTRODUCTION

The systematic review of data from multiple research studies is often required to answer many biomedical questions. The use of automated software tools to assist in reviewing research papers is a topic of increasing interest (1–7). Several research studies have described the use of LLMs, primarily the GPT-4.0 API or ChatGPT, to screen papers for specific criteria and for summarizing their content (8–14).

We previously assessed the use of GPT-4 to answer questions about studies on HIV drug resistance (HIVDR) (15). In that study, we found that GPT-4 reproducibly answered a set of 60 questions with a precision of 87% and a recall of 73% without human feedback. However, its performance was not improved with a 2000-word instruction sheet. The lack of improvement with this form of prompt engineering, led us to assess the degree to which fine-tuning could improve the performance of an LLM to specific answer questions about published HIVDR research studies.

We selected questions designed to determine whether a study reported HIV sequences and whether the sequences and their associated data were made publicly available. A fine-tuned model capable of answering questions about viral sequences, their public availability, and the demographics and antiviral treatments of the persons from whom the sequenced viruses were obtained would be invaluable to virology researchers, journal editors, and funding agencies.

## MATERIALS AND METHODS

### Fine-tuning

Figure 1 outlines the approach to fine-tuning, testing, and analysis used in this study. We worked with three LLMs: (1) GPT-4o mini-2024-07-18 (GPT-4o); (2) Meta-Llama 3.1-70B-Instruct (Llama3.1-70B); and (3) Meta-Llama 3.1-8B-Instruct (Llama3.1-8B). Llama3.1-70B and Llama3.1-8B have 70 billion and 8 billion parameters, respectively. The exact parameter count for GPT-4o has not been reported.

**Figure 1.**
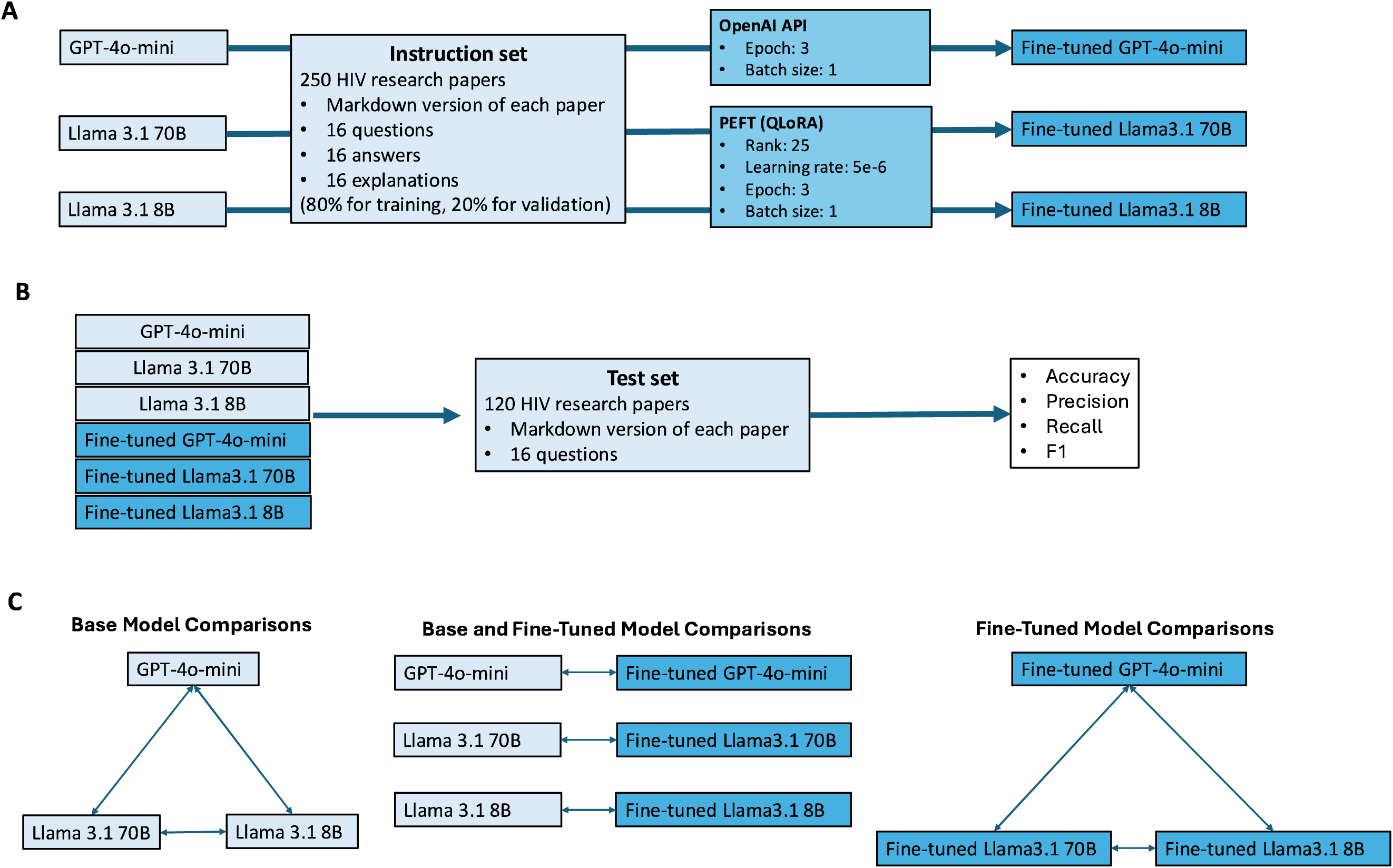
Approach to fine-tuning (A), testing (B), and analyses (C) performed in this study. Fine-tuning was performed using an instruction set comprising 250 marked-down research studies, 16 questions about each study, answers to each question, and explanations for each answer. GPT-4o was fine-tuned using the OpenAI API; Llama3.1-70B and Llama3.1-8B were fine-tuned using QLoRA (A). The accuracy, precision, recall, and F1-score of each base and fine-tuned model was assessed using a test set comprising 16 questions applied to 120 different published research studies on HIV drug resistance (B). Parametric (paired t-tests) and nonparametric (Wilcoxon signed-rank tests) methods were used to compare the performance of the base models to one another, the fine-tuned models to their respective base model, and the fine-tuned models to one another (C).

#### Research papers

We selected 250 curated research papers about HIV drug resistance from the Stanford HIV Drug Resistance Database (HIVDB) encompassing studies of (1) HIV sequences from infected persons who were either antiretroviral treatment (ART)-experienced or ART-naïve; (2) HIV isolates with known mutations undergoing *in vitro* susceptibility testing; (3) wildtype HIV isolates cultured in the presence of increasing concentrations of an antiretroviral drug (i.e., *in vitro* selection experiments); and (4) different approaches to HIV sequencing and cloning. The complete list of papers are in Supplementary Table 1.

#### Research questions

We designed 16 questions addressing key aspects of HIVDR including (1) Whether sequencing was performed on HIV isolates obtained from patients and whether the sequences were made publicly available (5 questions); (2) The demographics of patients whose viruses were sequenced (2 questions); (3) The treatment characteristics of patients whose viruses were sequenced (5 questions); and (4) The technical aspects of sequencing (4 questions). For eight questions, the answer was a list of items; for seven questions the answer was yes or no; and for one question, the answer was a number. For studies in which sequencing was not performed, the answers to patient demographics, treatments, and technical aspects of sequencing were considered to be “not reported”. Table 2 presents the complete list of questions along with their frequencies of being classified as true (for Boolean questions), non-empty (for list questions), or non-zero (for the single numeric question) in both the 250 study instruction set and the 120 study test set.

**Table 1.** GPU, VRAM, and Time Requirements Associated with Fine-Tuning and Testing.

|  | Fine-Tuning |  |  | Testing the Base Model |  |  | Testing the Fine-Tuned Model |  |  |
| --- | --- | --- | --- | --- | --- | --- | --- | --- | --- |
| <u>Model</u> | <u>GPU</u> | <u>VRAM</u> | <u>Time</u> | <u>GPU</u> | <u>VRAM</u> | <u>Time</u> | <u>GPU</u> | <u>VRAM</u> | <u>Time</u> |
| GPT-4o-mini | NA | NA | 2h | NA | NA | 1h | NA | NA | 1h |
| Llama3.1 8B | 1 A100 | 80G | 1h | 1 A100 | 80G | 2h | 2 A100 | 160G | 13h |
| Llama3.1 70B | 3 A100 | 240G | 7h | 3 A100 | 240G | 5h | 4 A100 | 320G | 21h |

Footnote: GPU (graphical processing unit); VRAM (video random access memory); A100 (Nvidia A100 tensor core GPU); VRAM is indicated as gigabytes.

**Table 2.** Complete List of Questions with their Frequencies of True, Non-Empty or Non-Zero in Both Instruction Set and Test Set.

|  | Question | Subject | Type | Instruction set (%) | Test set (%) |
| --- | --- | --- | --- | --- | --- |
| Q1 | Does the paper report HIV sequences from patient samples? | Data availability | Boolean | 85.6 | 66.7 |
| Q2 | Does the paper report in vitro drug susceptibility data? | Data availability | Boolean | 20 | 20.8 |
| Q3 | Were sequences from the paper made publicly available? | Data availability | Boolean | 56.4 | 17.5 |
| Q4 | What were the GenBank accession numbers for sequenced HIV isolates? | Data availability | List | 54.4 | 12.5 |
| Q5 | How many individuals had samples obtained for HIV sequencing? | Data availability | Number | 82.8 | 64.2 |
| Q6 | From which countries were the sequenced samples obtained? | Demographics | List | 76.8 | 56.7 |
| Q7 | From what years were the sequenced samples obtained? | Demographics | List | 64 | 51.7 |
| Q8 | Were samples cloned prior to sequencing? | Technical | Boolean | 2.8 | 2.5 |
| Q9 | Which HIV genes were reported to have been sequenced? | Technical | List | 91.2 | 75.0 |
| Q10 | What method was used for sequencing? | Technical | List | 64.8 | 45.8 |
| Q11 | What type of samples were sequenced? | Technical | List | 79.2 | 52.5 |
| Q12 | Were any sequences obtained from individuals with virological failure on a treatment regimen? | Treatment | Boolean | 36.4 | 30.8 |
| Q13 | Were the patients in the study in a clinical trial? | Treatment | Boolean | 14.4 | 15.8 |
| Q14 | Does the paper report HIV sequences from individuals who had previously received ARV drugs? | Treatment | Boolean | 46.4 | 46.7 |
| Q15 | Which drug classes were received by individuals in the study before sample sequencing? | Treatment | List | 36.8 | 38.3 |
| Q16 | Which drugs were received by individuals in the study before sample sequencing? | Treatment | List | 34.4 | 32.5 |

#### Instruction set

The instruction set contained 250 training samples. Each sample contained (1) a markdown version of one of the 250 papers containing its abstract, methods, results, discussion, and data sharing statement; (2) the 16 research questions; (3) the answers to the research questions; and (4) the explanation for each of the answers, including the text relevant to each answer. For questions not addressed by a study, the explanation indicated that the study did not address the question. The complete training set is in Supplementary Table 2.

#### Training hyperparameters

We used Hugging Face’s parameter efficient fine-tuning (16) using Quantized Low-Rank Adaptation (QLoRA) (17,18). Because our dataset was complex, we used a rank of 25, which is in the upper range of the recommended values of 4 to 32. We set our batch size to one because our sample sizes were large with the median number of tokens per sample being 9343 (range: 4261-22085).

Implementation.

Table 1 shows the GPU, VRAM, and time requirements associated with fine-tuning and testing each model used in this study. For GPT-4o, the GPU and VRAM requirements were not known because fine-tuning and testing were done using the OpenAI API (19).

### Testing and analysis

We created a test set comprising 120 journal articles. One hundred studies were identified by querying PubMed for journal articles about HIV drug resistance published in 2023. Twenty additional studies were selected from HIVDB because they reported data on uncommon topics that were unlikely to be reported in the first 100 studies. Like the training set, these papers included studies of viral sequences from HIV-infected persons, *in vitro* susceptibility testing, *in vitro* selection experiments, and technical aspects of HIV sequencing. Supplementary Table 3 lists the 120 papers used for testing.

For each question, we evaluated the answers of the original and fine-tuned models. Model-generated answers were compared to the human-curated answers, which were considered to be the ground truth. For the seven Boolean questions, we calculated the number of true positives, true negatives, false positives, and false negatives, as well as the model’s precision, recall, accuracy, and F1-score. For the eight list-based questions, we defined true positive when the model outputted a non-empty list exactly matching the human answer; true negative when both the human answer and the model output were empty lists; false positive when the human answer was an empty list while the model outputted a non-empty list; and false negative when the human answer was a non-empty list, but the model output was either an empty list or a list that did not match the human list. A similar approach was applied to the sole numeric question in that a result of 0 was considered analogous to an empty list. Supplementary Table 4 lists the correct answers and the answers for the three base and fine-tuned models for 1920 questions (120 papers x 16 questions).

We used Fisher Exact Tests to compare the accuracy, recall, and precision of the base model and fine-tuned model on the individual questions. For these tests, a Benjamini-Hochberg adjustment was calculated for the 16 questions evaluated. We used parametric (paired T-tests) and nonparametric (Wilcoxon-Rank Sign Test) tests to compare the average accuracy, recall, precision, and F1-score across questions of (1) the base models to one another; (2) the fine-tuned models to one another; and (3) each fine-tuned model to its base model. For these nine comparison, a Benjamini-Hochberg adjustment was calculated. A summary of the statistical analysis is in Supplementary File 5.

## RESULTS

Figure 2 displays the accuracy, precision, recall, and F1-score of the base and fine-tuned models for each of the 16 questions, individually. Points to the upper left of the diagonal line indicate questions for which there was any improvement for the fine-tuned model compared with the base model. Table 3 displays the accuracy, precision, recall, and F1-scores for the base and fine-tuned GPT-4o and Llama3.1-70B models for those questions for which there was a significant increase in either precision or recall for the fine-tuned model by Fisher Exact testing. For GPT-4o, the questions with improvements were Q2, Q6, Q9, Q11, Q14, Q15, and Q16. For Llama-70B, the questions with improvements were Q14, Q15, and Q16. The fine-tuning of Llama3.1-8B did not result in a significant increase in precision or recall for any question.

**Figure 2.**
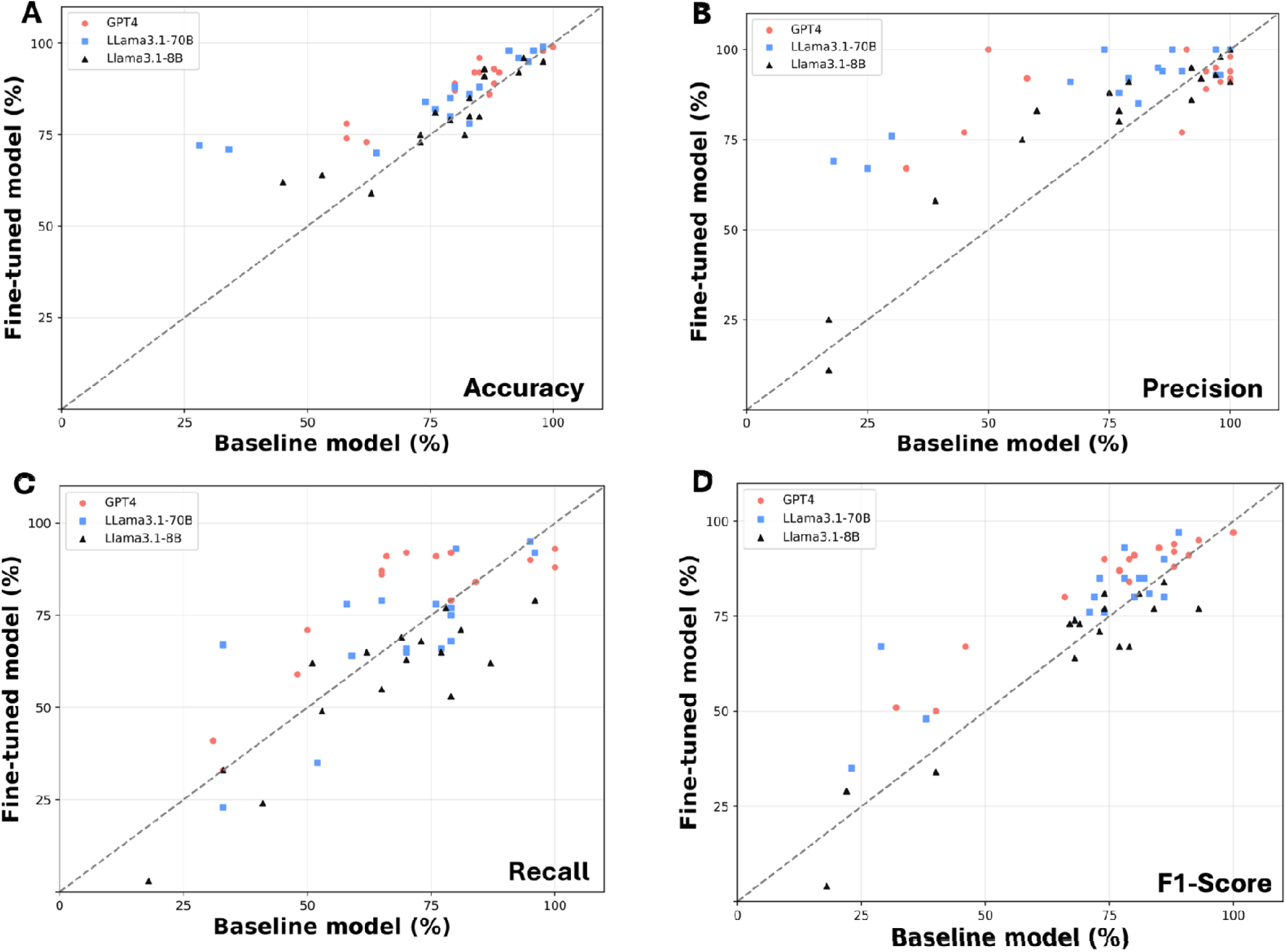
Comparison of base and fine-tuned models for each of the 16 questions applied to 120 published test studies. The accuracy (A), precision (B), recall (C), and F1-score (D) of the GPT4o, Llama3.1-70B, and Llama3.1-8B models are shown with the metrics for the base model indicated on the X-axis and for the fine-tuned model indicated on the Y-axis. Points to the left of the diagonal line indicate those questions for which there was an improvement for the fine-tuned model compared with the base model.

**Table 3.**
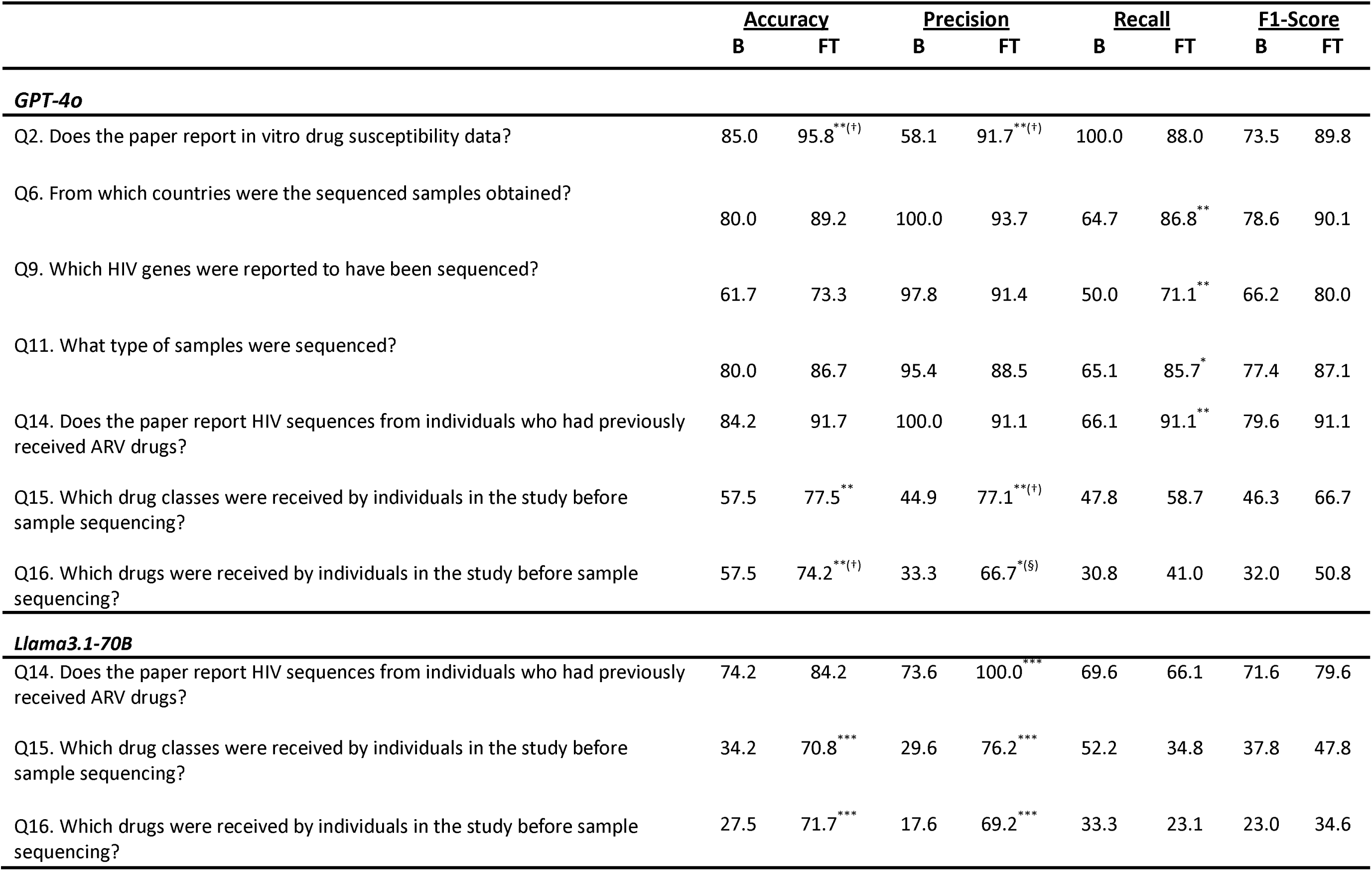
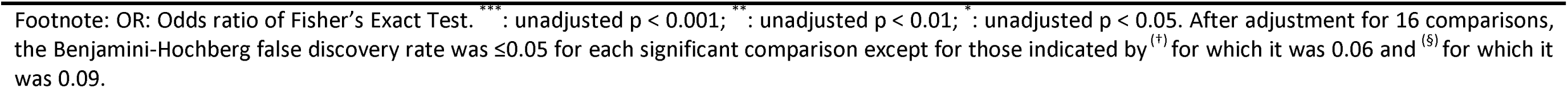
Accuracy, Precision, Recall and F1 Score for the Research Questions for which an Improvement was Observed After Fine-Tuning (FT) of Either GPT-4o or Llama3.1-70B.

|  | <u>Accuracy</u> |  | <u>Precision</u> |  | <u>Recall</u> |  | <u>F1-Score</u> |  |
| --- | --- | --- | --- | --- | --- | --- | --- | --- |
|  | B | FT | B | FT | B | FT | B | FT |
| ***GPT-4o*** |  |  |  |  |  |  |  |  |
| Q2. Does the paper report in vitro drug susceptibility data? | 85.0 | 95.8<sup>**(+)</sup> | 58.1 | 91.7<sup>**(+)</sup> | 100.0 | 88.0 | 73.5 | 89.8 |
| Q6. From which countries were the sequenced samples obtained? | 80.0 | 89.2 | 100.0 | 93.7 | 64.7 | 86.8<sup>**</sup> | 78.6 | 90.1 |
| Q9. Which HIV genes were reported to have been sequenced? | 61.7 | 73.3 | 97.8 | 91.4 | 50.0 | 71.1<sup>**</sup> | 66.2 | 80.0 |
| Q11. What type of samples were sequenced? | 80.0 | 86.7 | 95.4 | 88.5 | 65.1 | 85.7<sup>*</sup> | 77.4 | 87.1 |
| Q14. Does the paper report HIV sequences from individuals who had previously received ARV drugs? | 84.2 | 91.7 | 100.0 | 91.1 | 66.1 | 91.1<sup>**</sup> | 79.6 | 91.1 |
| Q15. Which drug classes were received by individuals in the study before sample sequencing? | 57.5 | 77.5<sup>**</sup> | 44.9 | 77.1<sup>**(+)</sup> | 47.8 | 58.7 | 46.3 | 66.7 |
| Q16. Which drugs were received by individuals in the study before sample sequencing? | 57.5 | 74.2<sup>**(+)</sup> | 33.3 | 66.7<sup>*(§)</sup> | 30.8 | 41.0 | 32.0 | 50.8 |
| ***Llama3.1-70B*** |  |  |  |  |  |  |  |  |
| Q14. Does the paper report HIV sequences from individuals who had previously received ARV drugs? | 74.2 | 84.2 | 73.6 | 100.0<sup>***</sup> | 69.6 | 66.1 | 71.6 | 79.6 |
| Q15. Which drug classes were received by individuals in the study before sample sequencing? | 34.2 | 70.8<sup>***</sup> | 29.6 | 76.2<sup>***</sup> | 52.2 | 34.8 | 37.8 | 47.8 |
| Q16. Which drugs were received by individuals in the study before sample sequencing? | 27.5 | 71.7<sup>***</sup> | 17.6 | 69.2<sup>***</sup> | 33.3 | 23.1 | 23.0 | 34.6 |

Footnote: OR: Odds ratio of Fisher's Exact Test. \*\*\* : unadjusted $p < 0.001$ ; \*\* : unadjusted $p < 0.01$ ; \* : unadjusted $p < 0.05$ . After adjustment for 16 comparisons, the Benjamini-Hochberg false discovery rate was $\leq 0.05$ for each significant comparison except for those indicated by <sup>(†)</sup> for which it was 0.06 and <sup>(§)</sup> for which it was 0.09.

Figure 3 compares the overall mean accuracy, precision, recall, and F1-score for the 16 questions pooled over the 120 test set studies. Prior to fine-tuning, GPT-4o displayed significantly greater precision and recall compared with Llama3.1-8B and significantly greater precision compared with Llama3.1-70B using both paired t-tests and Wilcoxon-ranked sign tests (Figure 2A). There were no statistically significant differences between Llama3.1-70B and Llama3.1-8B base models.

**Figure 3.**
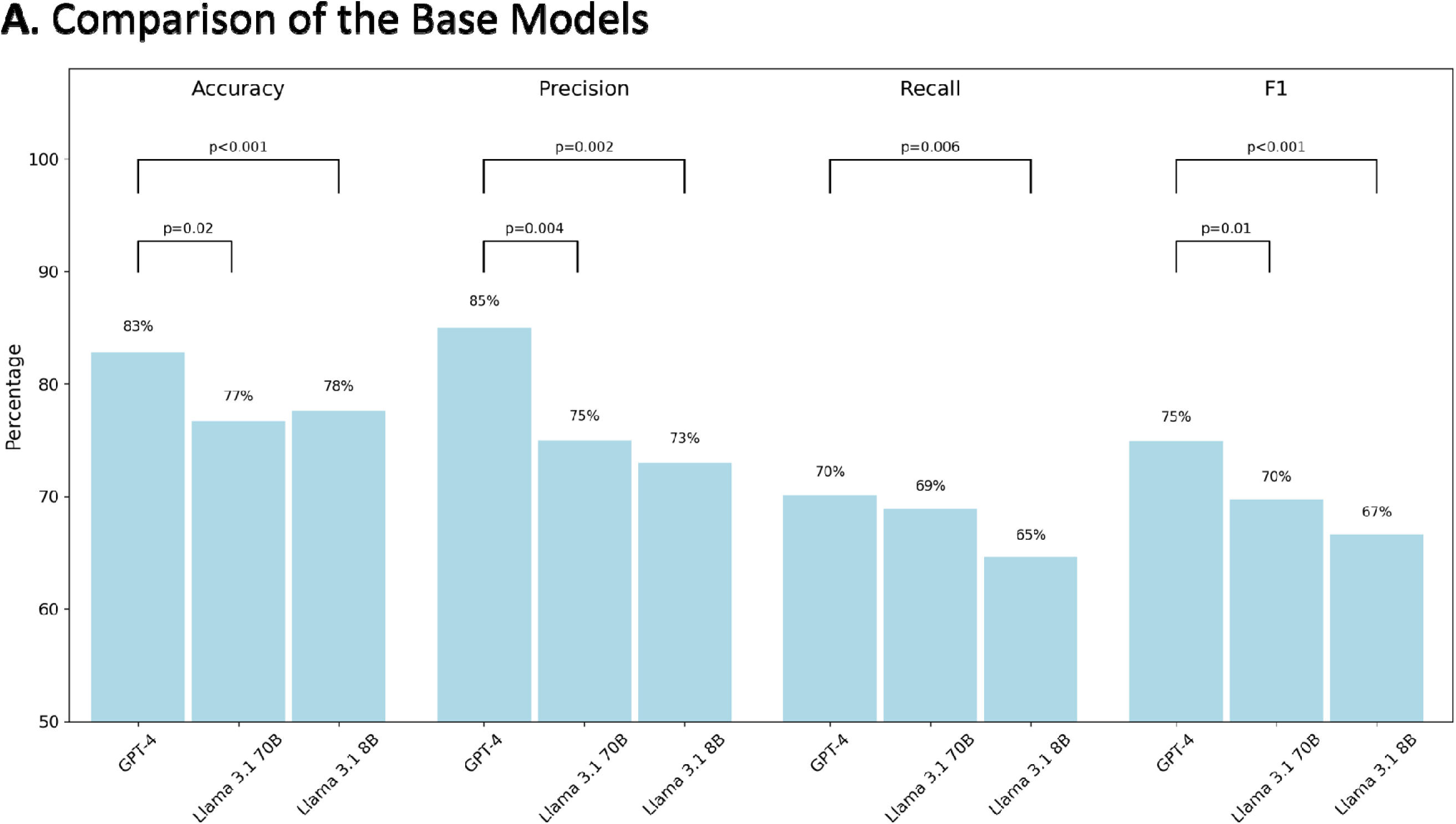
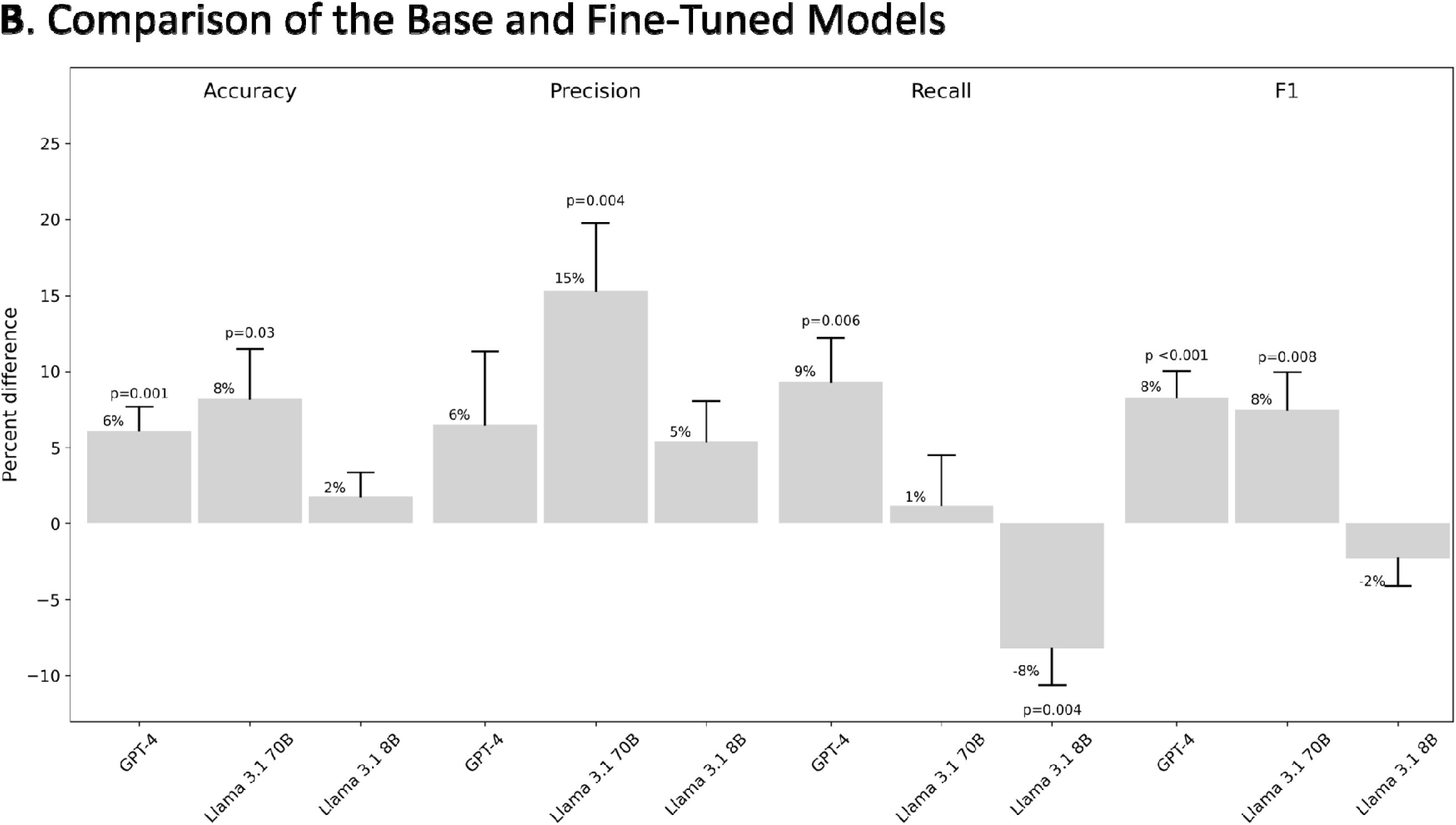
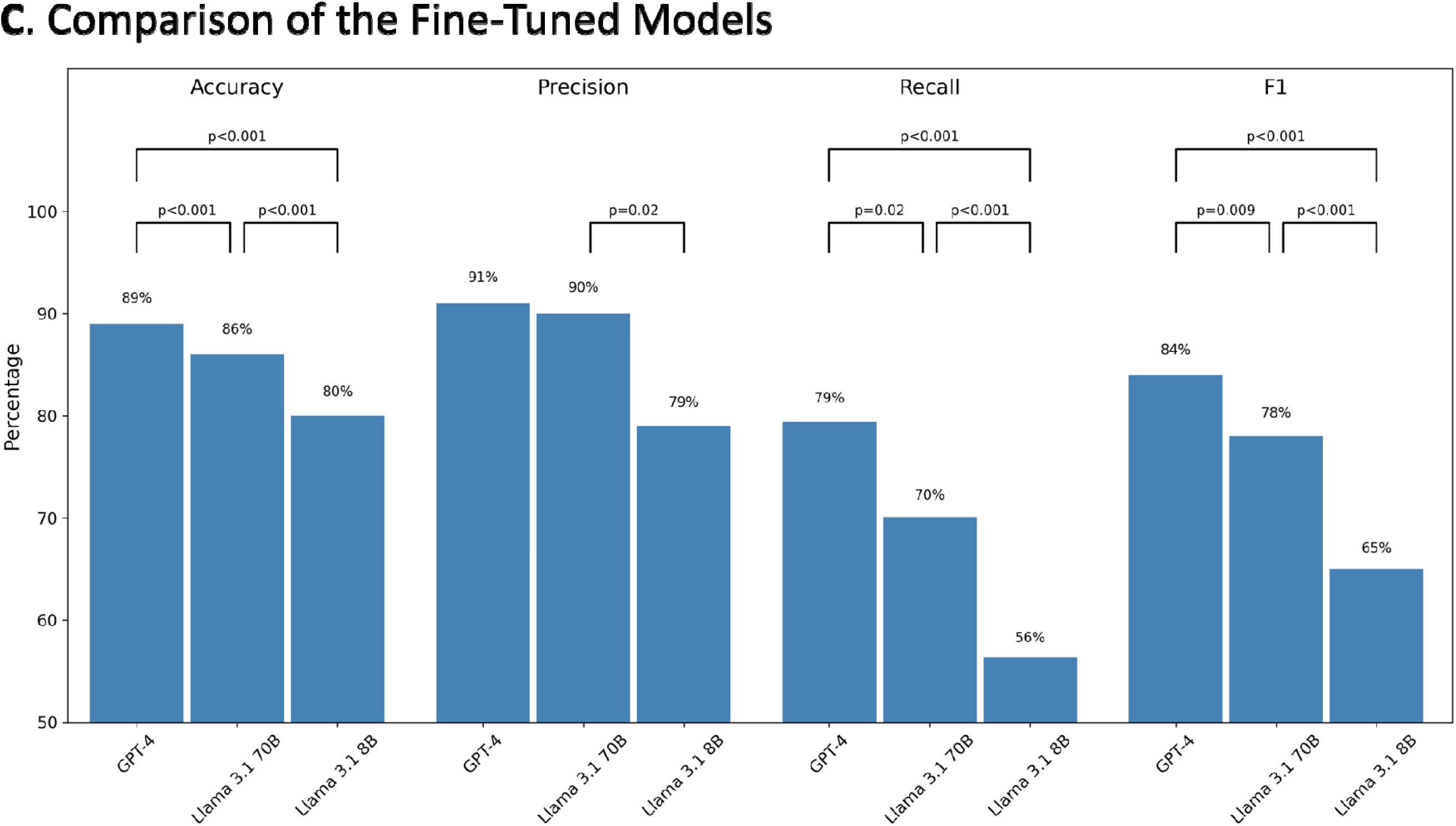
Comparisons of the performance of the base models to one another (A), the fine-tuned models to their respective base model (B), and the fine-tuned models to one another (C). The histograms in figures 3A and 3C represent the performance of the base and fine-tuned models, respectively. The histograms in figure 3B represent the difference in performance between the fine-tuned and base model. The error bars in figure 3B represent the standard error of the mean of the paired differences between the fine-tuned and base models. The mean differences in accuracy, precision, recall, and F1-score between models are indicated above the relevant histograms when statistically significant using both parametric (paired t-test) and nonparametric (Wilcoxon signed-rank test) methods. The p values shown are for the paired t-test performed on the aggregate data for each of the 16 questions. After adjustment for nine comparisons, the Benjamini-Hochberg false discovery rate was ≤0.05 for each of the p values shown.

After fine-tuning, GPT-4o displayed 6% increased accuracy, 6% increased precision, 9% increased recall, and 8% increased F1-score (Figure 2B). Llama3.1-70B displayed 8% increased accuracy, 15% increased precision, 1% increased recall, and 8% increased F1-score. Llama3.1-8B did not display significantly improved performance after fine-tuning. The increased recall, accuracy, and F1-score for the fine-tuned GPT-4o model and the increased precision, accuracy, and F1-score for the Llama3.1-70B model were statistically significant using both paired t-tests and Wilcoxon-ranked sign tests.

Figure 2C shows that the fine-tuned GPT-4o model displayed significantly greater accuracy, recall, and F1-score compared with both the fine-tuned Llama3.1-8B and Llama3.1-70B models using both paired t-tests and Wilcoxon-ranked sign tests. Llama3.1-70B displayed significantly greater accuracy, recall, and F1-score compared with Llama3.1-8B using both paired t-tests and Wilcoxon-ranked sign tests.

## DISCUSSION

Fine-tuning a foundation model provides significant advantages for handling domain-specific tasks. By training a model on targeted data, it becomes more reliable and effective in delivering accurate results without requiring complex prompts. Using a pre-trained model for fine-tuning lowers computational costs making it a highly efficient approach for specialized use cases. This study demonstrates that fine-tuning GPT-4o and Llama3.1-70B significantly improved their ability to answer questions about research studies in a specialized medical field, specifically those questions included in their training.

Our findings can be distilled into four main observations: (1) Prior to fine-tuning, GPT-4o displayed significantly greater performance than both Llama3.1-70B and Llama3.1-8B as a result of increased precision compared with Llama3.1-70B and increased precision and recall compared with Llama3.1-8B, while no difference in performance between Llama3.1-70B and Llama3.1-8B was observed; (2) After fine-tuning both GPT-4o and Llama3.1-70B, but not Llama3.1-8B, displayed significantly improved performance compared with their base models; (3) The improved performance of GPT-4o was a result of its 6% increased precision and 9% increased recall while the improved performance of Llama3.1-70B resulted from its 15% increased precision; (4) After fine-tuning, Llama3.1-70B outperformed Llama3.1-8B primarily as a result of its improved precision, but still did not perform as well as the fine-tuned GPT-4o model which displayed superior recall.

Most studies evaluating the potential use of LLMs for answering questions about research studies have evaluated the use of foundational models to determine whether the titles and abstracts of a study were likely to meet the inclusion criteria for a systematic review (8–14,20). Few have evaluated fine-tuned foundational models for their ability to answer questions about full-text research papers (21,22).

We examined the effects of fine-tuning on three models: GPT-4o, selected for its top-tier performance and ease of fine-tuning with just an API and Python script (23); Llama3.1-70B, chosen for its long context length and high ranking among open-source models (24); and Llama3.1-8B, to assess fine-tuning’s impact on a smaller model. We intended to fine-tune the even larger Llama3.1-405B model, but the most widely available GPUs lacked the memory capacity to train this model, despite several optimization attempts. Testing this larger model would have required even more memory, and renting the necessary GPUs was cost-prohibitive at significantly more than $10,000 for both fine-tuning and testing.

LoRA and QLoRA are widely used approaches for fine-tuning foundational models, as they adjust a small subset of parameters, reducing both memory usage and computational costs compared to full fine-tuning (17). LoRA adapters can be reused to fine-tune multiple foundational models and enable low-cost re-tuning when these models are updated. Moreover, LoRA adapters can be easily swapped or combined, facilitating modular specialization (17). QLoRA further introduces innovations that optimize memory usage while maintaining performance (18).

We selected a narrow topic to determine whether a foundation model could be fine-tuned to answer questions about full-text research studies. Given the specificity of our topic, we chose not to expand our training set or further optimize our model. However, the questions able to be answered by the fine-tuned models target key data types broadly relevant to studies of pathogenic human viruses, including those with available antiviral treatments, as well as those with pandemic potential. Therefore, the success that we have described is a step towards accomplishing the more ambitious goal of developing a fine-tuned model capable of answering questions broadly applicable to all pathogenic human viruses.

## Data availability statement

All data generated or analyzed during this study are included in this published article and its supplementary information files. The Llama3.1-70B and Llama3.1-8B LoRA adapters developed for this study was shared on the Hugging Face platform (https://huggingface.co/kmtao/llama3.1-8B-HIVDB-adapter, https://huggingface.co/kmtao/llama3.1-70B-HIVDB-adapter).

## Author contributions

Conceptualization: K.T., V.A., and R.W.S; Data Curation: K.T, J.Z and Z.A.O; Formal analysis: K.T, J.Z, C.S and R.W.S; Methodology: K.T, J.Z, J.A and R.W.S; Writing – Original Draft Preparation: K.T, J.Z and R.W.S; Writing – Review & Editing: K.T, J.Z, V.A, C.S., and R.W.S

## SUPPORT INFORMATION CAPTIONS

<u>Supplementary Table 1 (S1Table.xlsx)</u>: The list of research studies used for the instruction set.

<u>Supplementary Table 2 (S2Table.xlsx)</u>: The list of training examples included in the instruction set.

<u>Supplementary Table 3 (S3Table.xlsx)</u>: The list of research studies used for testing.

<u>Supplementary Table 4 (S4Table.xlsx)</u>: The correct answers and the answers of each of the six models (base and fine-tuned for GPT-4o, Llama3.1-70B, and Llama3.1-8B) for 1920 questions (120 test studies x 16 questions).

<u>Supplementary File 5 (S5Appendix.xlsx)</u>: Tab 1 contains the raw data and results of Fisher Exact Tests for each of the 16 questions for each of the six models (base and fine-tuned for GPT-4o, Llama3.1-70B, and Llama3.1-8B). Tab 2 contains the raw data and nine comparisons between the models. Specifically, paired t-tests and Wilcoxon signed-rank tests were used to compare the base models to one another, the fine-tuned models to their respective base model, and the fine-tuned models to one another. Tab 3 illustrates how the Benjamini-Hochberg adjustment for the nine model comparisons was performed.

